# CarotidMamba: Foundation Model–Enabled CTA Phenotyping of Symptomatic Carotid Plaques in a Multi-Center Retrospective Study

**DOI:** 10.64898/2026.06.02.26354776

**Authors:** Yong-Sheng Liu, Xin-Wei Dou, Peng-Yu Zheng, Wang Feng, Liu-Jie Ma, Ying-Ning You, Gui-Wen Shao, Jia-Geng Shen, Xin Yu, Chen Qiao, Zi-Wei Cheng, Zhong-Wen Li, Feng Su, Bo-Wen Zhang, Xing-Huang Qu, Gui-Nan Jiang

## Abstract

**Background:** Treatment decisions for carotid atherosclerotic disease rely primarily on luminal stenosis, although plaque vulnerability and symptomatic status better reflect short-term cerebrovascular risk. A scalable CTA tool for automated phenotyping of symptomatic carotid disease is lacking.

**Materials & Methods:** In this multi-institutional retrospective study, 689 patients (mean age, 67.9 ± 7.7 years; 366 men) from four hospitals were analyzed after screening 705 CTA examinations. 423 patients from one center were used for five-fold development and internal validation, and 266 patients from three centers for independent external validation. CarotidMamba, a deep learning framework combining dual foundation-model encoders with Mamba-based sequence modeling, was developed and benchmarked against clinical, radiomics, clinic-radiomics, CNN, and transformer comparators.

**Results:** In the development cohort, CarotidMamba achieved an AUC of 0.839 (95% CI, 0.799–0.879) and accuracy of 0.825 (95% CI, 0.793–0.857), outperforming the strongest comparator by 0.066 and 0.050, respectively. External validation yielded AUCs of 0.897 (95% CI, 0.835–0.959) in YCH, 0.809 (95% CI, 0.720–0.898) in DCH, and 0.762 (95% CI, 0.649–0.875) in GH-NTC. CarotidMamba showed the lowest Brier score and expected calibration error across cohorts, with calibration slopes near 1.0.

**Conclusion:** CarotidMamba provides an interpretable, clinically oriented, and externally validated CTA framework for phenotyping symptomatic carotid plaques, supporting vulnerability-aware imaging assessment beyond stenosis alone.

## Introduction

Stroke remains a leading cause of death and disability worldwide, and carotid atherosclerotic disease accounts for a substantial fraction of ischemic cerebrovascular events [1,2]. For decades, treatment selection for carotid disease has been anchored to stenosis severity thresholds, with revascularization generally considered for symptomatic stenosis of 50% or greater and for selected asymptomatic stenosis above 60% [3]. Yet plaque biology is not fully captured by lumen narrowing alone. Intraplaque hemorrhage, lipid-rich necrotic core, ulceration, and related vulnerability features are more directly linked to ischemic risk, recurrent events, and treatment benefit than stenosis severity by itself [4–8]. As medical therapy has improved and the role of routine carotid revascularization has narrowed, identifying the subgroup of patients who truly harbor high-risk disease has become increasingly important [8,9].

Computed tomography angiography (CTA) is widely available in both acute and elective workflows, offers high spatial resolution, and can characterize plaque morphology, calcification patterns, ulceration, and other imaging biomarkers relevant to vulnerability [5,6,10,11]. However, manual CTA assessment is time-consuming, observer-dependent, and difficult to scale. This has motivated artificial intelligence (AI) approaches for automated carotid plaque analysis. Existing CTA studies suggest that clinical models, radiomics, and clinic-radiomics models can identify symptomatic or otherwise high-risk plaques beyond conventional visual assessment [12,15–20]. Nevertheless, these approaches often depend on manual or semi-manual region-of-interest delineation, carefully engineered handcrafted features, and relatively narrow single-center data distributions, which can limit reproducibility and generalizability [10–12,15–20].

Deep learning offers a route to more automated CTA phenotyping [13,14], but important challenges remain. Conventional convolutional neural networks (CNNs) are effective local feature extractors yet may not fully capture the long-range spatial context contained within an entire carotid CTA examination. Transformer-based models can model broader dependencies, but their quadratic sequence complexity can become burdensome for long ordered scan sequences and may encourage overfitting in moderate-sized clinical datasets [29]. At the same time, medical foundation models provide transferable priors that are difficult to learn from a single vascular dataset alone. MedCLIP captures disease-relevant medical semantics from image–text contrastive learning [21], whereas SAM and its medical variant SAM-Med2D provide strong anatomy-aware priors inherited from large-scale segmentation pretraining [22,23]. Efficient state-space models, including Mamba, further offer a compelling way to model long ordered image sequences with linear complexity [24–26].

In this study, we developed CarotidMamba, a foundation model-enabled CTA analysis framework for classifying symptomatic versus asymptomatic carotid atherosclerotic disease. CarotidMamba combines dual foundation-model encoders with carotid-specific Mamba-based sequence modeling, enabling patient-level CTA phenotyping without lesion-level manual annotation during model training. In a retrospective multi-institutional cohort of 689 patients from four hospitals, we benchmarked CarotidMamba against clinical, radiomics, clinic-radiomics, CNN, and transformer baselines, and evaluated discrimination, calibration, decision-curve utility, and interpretability. We hypothesized that this design would improve clinically meaningful CTA phenotyping beyond existing paradigms.

## Results

### Clinical context and study cohorts

Figure 1a illustrates the clinical problem addressed in this study: discriminating symptomatic from asymptomatic carotid atherosclerotic disease on CTA, with symptomatic status used as a clinically pragmatic surrogate of plaque vulnerability. This framing aligns with the growing shift from stenosis-only assessment toward vulnerability-oriented carotid imaging.

**Figure 1.**
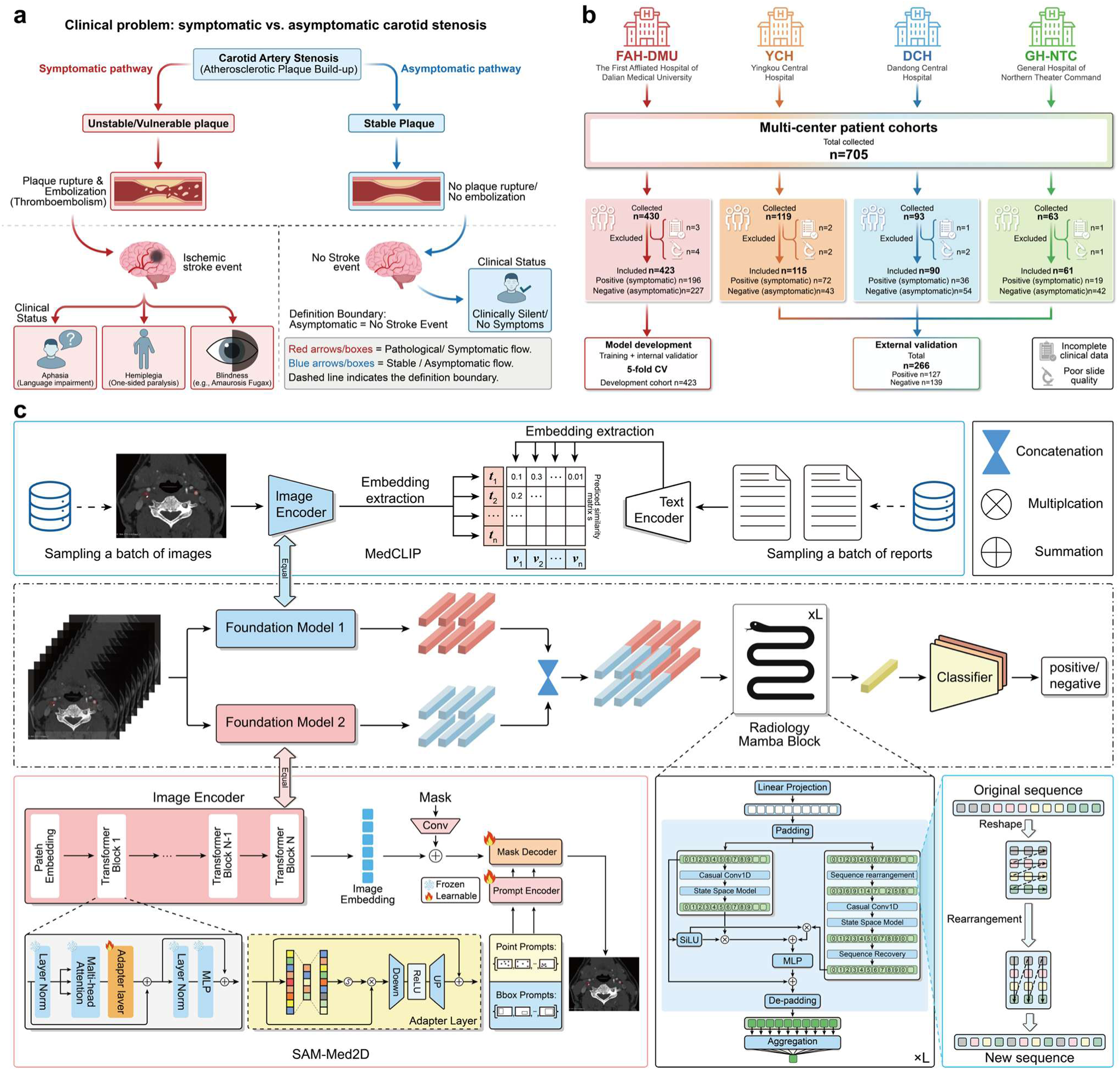
Clinical context, cohort construction, and the CarotidMamba framework. a, Conceptual illustration of symptomatic versus asymptomatic carotid atherosclerotic disease, emphasizing unstable plaque rupture and embolization in symptomatic disease and more stable plaque behavior in asymptomatic disease. b, Multicenter cohort assembly. A total of 705 CTA examinations were screened and 689 patients were included after exclusion for incomplete clinical data or poor image quality. FAH-DMU served as the development cohort, whereas YCH, DCH, and GH-NTC were used for independent external validation. c, Overview of CarotidMamba. Ordered CTA slices are passed through dual foundation encoders (MedCLIP and SAM-Med2D), fused by concatenation, processed by Radiology Mamba blocks with sequence rearrangement, pooled by attention, and classified as symptomatic or asymptomatic. Insets summarize the MedCLIP pretraining concept, the SAM-Med2D image encoder, and the internal structure of the Radiology Mamba block.

Across four participating centers, 705 CTA examinations were screened and 689 patients were retained after exclusion for incomplete clinical data or poor image quality (Fig. 1b and Table 1). The lead-center development cohort from the First Affiliated Hospital of Dalian Medical University (FAH-DMU) included 423 patients (196 symptomatic and 227 asymptomatic) and was used for stratified five-fold model development and internal validation. Three independent external cohorts contributed 115 patients from Yingkou Central Hospital (YCH), 90 from Dandong Central Hospital (DCH), and 61 from the General Hospital of Northern Theater Command (GH-NTC). In total, the analytic cohort comprised 323 symptomatic and 366 asymptomatic cases, with 266 patients reserved for fully independent external validation.

**Table 1.** Clinical characteristics of the development and external validation cohorts. Continuous variables are presented as mean ± s.d. and categorical variables as count (percentage). P values compare symptomatic and symptomatic patients within each center. DCH, Dandong Central Hospital; FAH-DMU, The First Affiliated Hospital of Dalian Medical University; GH-NTC, General Hospital of Northern Theater Command; YCH, Yingkou Central Hospital.

| Characteristic | FAH-DMU<br>sympto<br>matic | FAH-DMU<br>asympto<br>matic | FA<br>H-DM<br>U P | GH-NTC<br>sympto<br>matic | GH-NTC<br>asympto<br>matic | GH-NT<br>C P | DCH<br>sympto<br>matic | DCH<br>asympto<br>matic | DC<br>H P | YCH<br>sympto<br>matic | YCH<br>asympto<br>matic | YC<br>H P |
| --- | --- | --- | --- | --- | --- | --- | --- | --- | --- | --- | --- | --- |
| Age (years) | 68.40 ±<br>7.44 | 68.93 ±<br>7.56 | 0.62<br>0 | 68.20 ±<br>7.39 | 66.23 ±<br>7.47 | 0.43<br>1 | 69.19 ±<br>7.38 | 67.76 ±<br>7.58 | 0.31<br>2 | 67.38 ±<br>9.03 | 69.32 ±<br>8.23 | 0.28<br>1 |
| Gender, n (%) | 165<br>(84.2) | 179<br>(78.9) | 0.20<br>1 | 16 (84.2) | 32 (76.2) | 0.48<br>9 | 34 (94.4) | 48 (88.9) | 0.46<br>8 | 58 (80.6) | 28 (65.1) | 0.13<br>3 |
| Smoking, n (%) | 71 (36.2) | 61 (26.9) | 0.04<br>9 * | 10 (52.6) | 26 (61.9) | 0.71<br>6 | 16 (44.4) | 22 (40.7) | 0.89<br>6 | 38 (52.8) | 20 (46.5) | 0.61<br>6 |
| Alcohol, n (%) | 43 (22.1) | 40 (17.6) | 0.30<br>8 | 11 (57.9) | 21 (50.0) | 0.70<br>7 | 13 (36.1) | 13 (24.1) | 0.31<br>9 | 22 (30.6) | 14 (32.6) | 1.00<br>0 |
| Hypertension, n (%) | 130<br>(66.3) | 151<br>(67.1) | 0.94<br>7 | 9 (47.4) | 30 (71.4) | 0.14<br>4 | 21 (58.3) | 39 (72.2) | 0.25<br>4 | 57 (79.2) | 33 (76.7) | 1.00<br>0 |
| Hyperlipidemia, n<br>(%) | 69 (35.2) | 108<br>(48.0) | 0.01<br>1 * | 6 (31.6) | 15 (35.7) | 1.00<br>0 | 14 (40.0) | 26 (49.1) | 0.53<br>8 | 21 (29.2) | 15 (34.9) | 0.76<br>4 |
| Diabetes, n (%) | 71 (36.2) | 90 (39.8) | 0.51<br>0 | 5 (26.3) | 21 (50.0) | 0.14<br>3 | 12 (33.3) | 24 (44.4) | 0.40<br>4 | 22 (30.6) | 13 (30.2) | 1.00<br>0 |
| Heart disease, n (%) | 40 (20.4) | 41 (18.1) | 0.62<br>6 | 4 (21.1) | 14 (33.3) | 0.51<br>8 | 6 (17.1) | 11 (20.4) | 0.91<br>8 | 13 (18.1) | 14 (32.6) | 0.15<br>1 |
| Neurological<br>abnormality, n (%) | 11 (5.6) | 7 (3.1) | 0.29<br>7 | 10 (52.6) | 7 (16.7) | 0.03<br>9 * | 13 (36.1) | 11 (20.4) | 0.15<br>8 | 33 (45.8) | 10 (23.3) | 0.01<br>8 * |
| Urinary disease, n (%) | 4 (2.3) | 10 (4.8) | 0.27<br>6 | 4 (21.1) | 2 (4.8) | 0.09<br>0 | 2 (5.7) | 4 (7.4) | 1.00<br>0 | 4 (5.6) | 2 (4.7) | 1.00<br>0 |
| Lung disease, n (%) | 8 (4.1) | 6 (2.6) | 0.58<br>1 | 8 (42.1) | 17 (40.5) | 1.00<br>0 | 2 (5.7) | 1 (1.9) | 0.55<br>9 | 0 (0.0) | 0 (0.0) | 1.00<br>0 |
| Surgery history, n (%) | 45 (23.0) | 74 (32.6) | 0.03<br>7 * | 8 (42.1) | 21 (50.0) | 0.65<br>5 | 14 (40.0) | 23 (42.6) | 0.98<br>2 | 12 (16.7) | 6 (14.0) | 0.83<br>8 |
| Allergy history, n (%) | 5 (2.6) | 12 (5.3) | 0.23<br>8 | 1 (5.3) | 5 (11.9) | 0.67<br>0 | 2 (5.7) | 1 (1.9) | 0.55<br>9 | 3 (4.2) | 2 (4.7) | 1.00<br>0 |
| Elevated uric acid, n (%) | 17 (9.0) | 29 (13.3) | 0.23<br>3 | 1 (5.3) | 10 (23.8) | 0.25<br>9 | 22 (68.8) | 31 (58.5) | 0.47<br>5 | 4 (5.6) | 2 (4.7) | 1.00<br>0 |
| Abnormal ECG, n<br>(%) | 116<br>(62.7) | 128<br>(59.8) | 0.62<br>6 | 5 (26.3) | 15 (35.7) | 0.75<br>4 | 21 (70.0) | 45 (88.2) | 0.08<br>1 | 40 (55.6) | 17 (39.5) | 0.14<br>0 |
| Elevated<br>homocysteine, n (%) | 86 (43.9) | 68 (30.4) | 0.00<br>6 ** | 0 (0.0) | 1 (2.4) | 1.00<br>0 | 24 (80.0) | 39 (76.5) | 0.92<br>7 | 32 (44.4) | 18 (41.9) | 0.80<br>7 |

### Model development and study workflow

Structured clinical variables were first evaluated in the FAH-DMU development cohort and used to build a logistic-regression clinical model. In univariable analysis, smoking was positively associated with symptomatic status (odds ratio [OR], 1.55; 95% CI, 1.02–2.34; P = 0.039), whereas hyperlipidemia (OR, 0.59; 95% CI, 0.40–0.87; P = 0.008) and prior surgery history (OR, 0.62; 95% CI, 0.40–0.95; P = 0.029) were inversely associated; elevated homocysteine showed the strongest positive association (OR, 1.79; 95% CI, 1.20–2.68; P = 0.004). After multivariable adjustment, hyperlipidemia, prior surgery history, and elevated homocysteine remained independently associated with symptomatic status, whereas smoking became borderline (Table 1 and Fig. 2d). These analyses supported the clinical comparator model but also suggested that structured variables alone would not fully capture the symptomatic plaque phenotype. To reflect the main current CTA-analysis paradigms, we additionally constructed radiomics, clinic-radiomics, CNN, and transformer comparators using the same cohort partitions.

**Figure 2.**
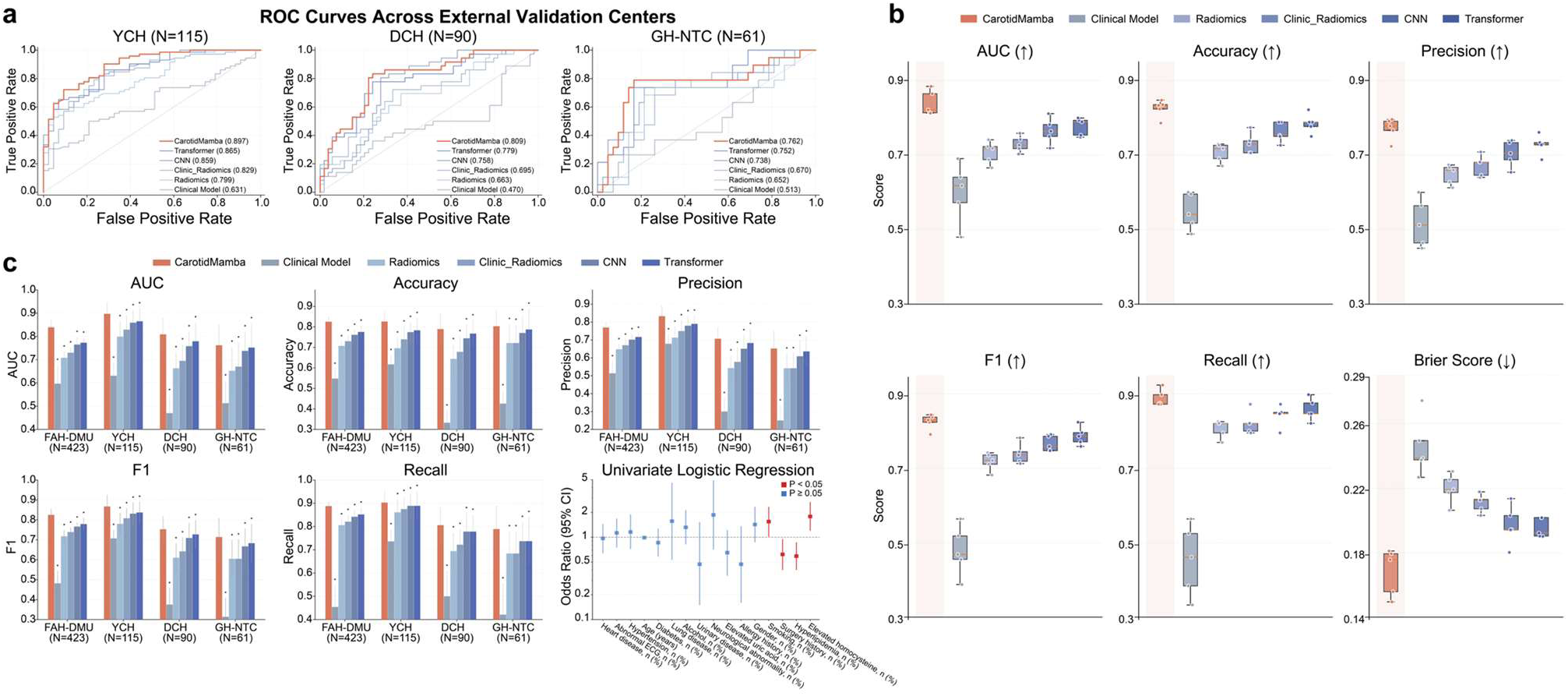
Discriminative performance of CarotidMamba compared with clinical, radiomics, and deep learning baselines. a, Receiver operating characteristic curves in the three external validation cohorts. b, Distribution of AUC, accuracy, precision, F1 score, recall, and Brier score across the tested methods. c, Cohort-level comparison of discrimination metrics across FAH-DMU, YCH, DCH, and GH-NTC. d, Development-cohort regression summary for selected structured clinical variables associated with symptomatic status.

The overall CarotidMamba workflow is summarized in Fig. 1c. Each CTA examination was represented as an ordered sequence of axial slices. Slice-level features were extracted in parallel from foundation models MedCLIP and SAM-Med2D, providing complementary disease-aware and anatomy-aware representations. These features were concatenated and passed to a carotid-specific Radiology Mamba encoder that included a sequence-rearrangement branch and attention pooling to generate the final patient-level prediction. This design aimed to preserve clinically relevant whole-scan context while remaining computationally efficient for long CTA sequences.

### CarotidMamba improves discrimination in internal and external validation

On five-fold development and internal validation in FAH-DMU, CarotidMamba achieved an AUC of 0.839 (95% CI, 0.799–0.879), accuracy of 0.825 (95% CI, 0.793–0.857), precision of 0.770 (95% CI, 0.736–0.804), F1 score of 0.825 (95% CI, 0.785–0.865), and recall of 0.888 (95% CI, 0.863–0.913). These values exceeded those of the clinical model, radiomics model, clinic-radiomics model, CNN baseline, and transformer baseline (Fig. 2b,c). Relative to the strongest comparator, the transformer model, CarotidMamba improved AUC by 0.066 and accuracy by 0.050, while also yielding higher precision, F1 score, and recall.

Performance remained robust under independent external validation. In YCH, CarotidMamba achieved an AUC of 0.897 (95% CI, 0.835–0.959), accuracy of 0.826 (95% CI, 0.761–0.891), F1 score of 0.867 (95% CI, 0.794–0.940), and recall of 0.903 (95% CI, 0.842–0.964). In DCH, the corresponding values were 0.809 (95% CI, 0.720–0.898), 0.789 (95% CI, 0.691–0.887), 0.753 (95% CI, 0.672–0.834), and 0.806 (95% CI, 0.712–0.900). In GH-NTC, they were 0.762 (95% CI, 0.649–0.875), 0.803 (95% CI, 0.701–0.905), 0.714 (95% CI, 0.596–0.832), and 0.789 (95% CI, 0.664–0.914), respectively (Fig. 2a,c). Across the three external cohorts, CarotidMamba improved AUC over the strongest comparator by 0.032 in YCH, 0.030 in DCH, and 0.010 in GH-NTC. Importantly, it also maintained high recall across cohorts, a desirable property for flagging symptomatic disease.

### Calibration, decision utility, and error patterns support clinical relevance

Discrimination alone is insufficient for clinical translation, so we next examined calibration and decision-curve behavior. As shown in Fig. 3a,b, CarotidMamba consistently produced the most favorable calibration profile across centers. It achieved the lowest Brier score in every cohort: 0.1596 in FAH-DMU, 0.1249 in YCH, 0.1769 in DCH, and 0.1724 in GH-NTC. Expected calibration error was likewise the lowest for CarotidMamba in each cohort, ranging from 0.0429 in YCH to 0.1759 in GH-NTC. Calibration slopes remained close to unity (0.98 in FAH-DMU, 1.10 in YCH, 0.985 in DCH, and 0.95 in GH-NTC), indicating that predicted probabilities were neither markedly overconfident nor underconfident.

**Figure 3.**
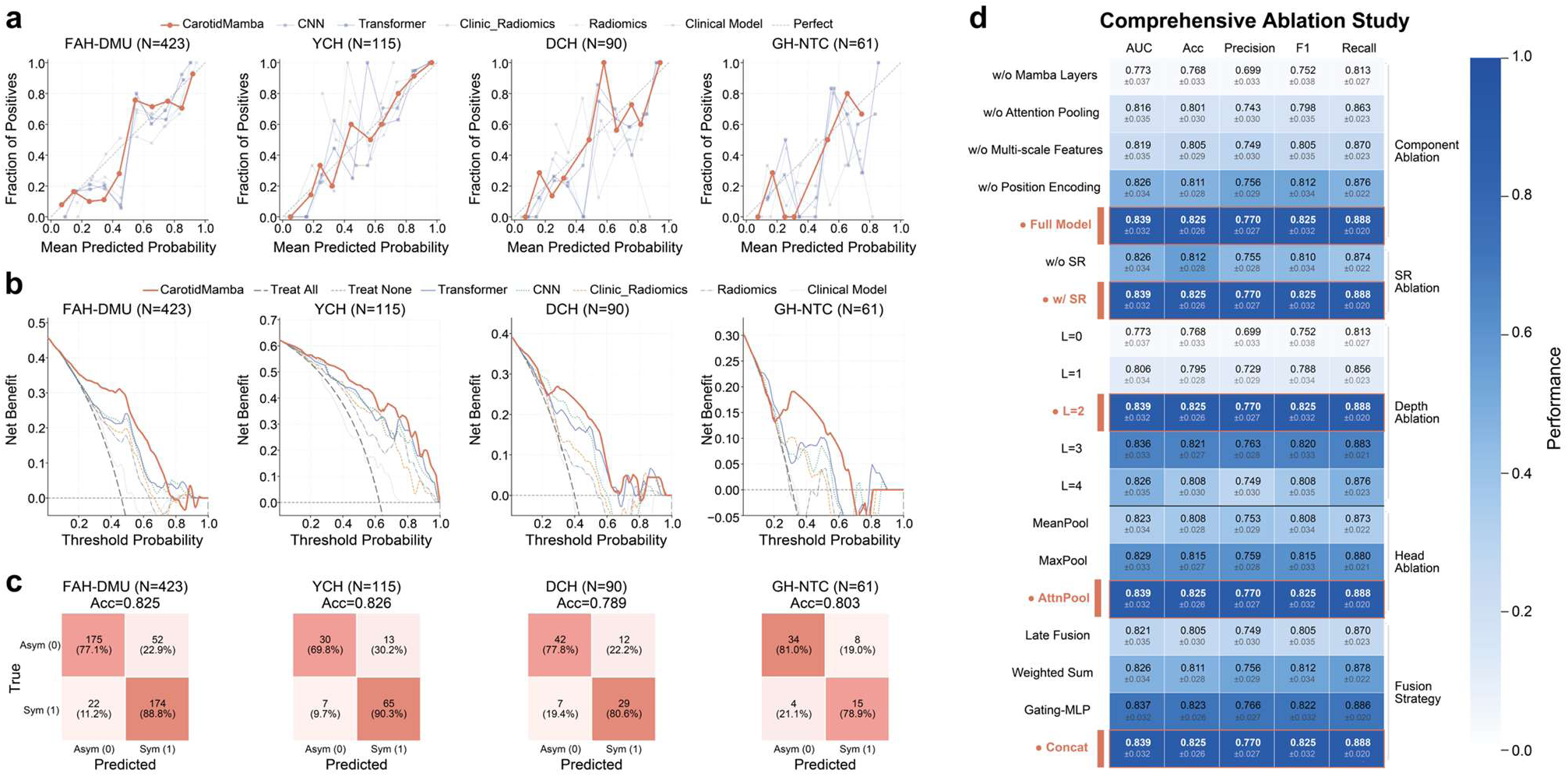
Calibration, decision utility, confusion matrices, and ablation analysis. a, Calibration curves of CarotidMamba and comparator methods in the development and external cohorts. b, Decision-curve analysis showing net benefit across threshold probabilities. c, Confusion matrices for CarotidMamba in each cohort. d, Comprehensive ablation map summarizing the effects of removing or modifying key architectural components, sequence rearrangement, encoder depth, pooling strategy, and fusion strategy.

Decision-curve analysis further showed that CarotidMamba yielded the highest net benefit over clinically relevant threshold ranges, with net benefit values of 0.3528 in FAH-DMU, 0.5652 in YCH, 0.2944 in DCH, and 0.1926 in GH-NTC. Confusion matrices (Fig. 3c) showed that these gains were not driven by a single class: symptomatic cases were captured with relatively few false negatives while asymptomatic cases remained reasonably well separated, supporting a balanced prediction profile rather than an extreme thresholding strategy.

### Dual foundation initialization and sequence aggregation underpin performance

Comprehensive ablation analysis showed that CarotidMamba depends on both representation quality and sequence modeling (Fig. 3d). Removing the Mamba layers caused the largest performance drop, reducing AUC from 0.839 (95% CI, 0.799–0.879) to 0.773 (95% CI, 0.727–0.819) and recall from 0.888 (95% CI, 0.863–0.913) to 0.813 (95% CI, 0.779–0.847). Removing attention pooling reduced AUC to 0.816 (95% CI, 0.773–0.859), removing multi-scale dual-encoder features reduced AUC to 0.819 (95% CI, 0.776–0.862), and removing positional encoding reduced AUC to 0.826 (95% CI, 0.784–0.868). Disabling sequence rearrangement also reduced AUC to 0.826 (95% CI, 0.784–0.868), indicating that the reordered branch contributed information beyond the native slice order.

Model depth mattered but showed diminishing returns. A two-block Mamba encoder performed best, slightly outperforming a three-block encoder (AUC, 0.836; 95% CI, 0.795–0.877), whereas both shallower and deeper variants underperformed. Attention pooling was preferable to mean pooling (AUC, 0.823; 95% CI, 0.781–0.865) and max pooling (AUC, 0.829; 95% CI, 0.788–0.870). Among fusion strategies, simple concatenation was strongest (AUC, 0.839; 95% CI, 0.799–0.879), slightly ahead of gating-MLP fusion (AUC, 0.837; 95% CI, 0.797–0.877) and clearly better than weighted-sum or late-fusion alternatives.

### Interpretability analyses support biological plausibility

Representative attention maps showed that CarotidMamba concentrated on plaque-bearing carotid territories rather than diffuse background structures (Fig. 4a). Across axial and reformatted views, high-response regions clustered around the carotid bifurcations and vessel-wall interfaces, where plaque morphology would be expected to be most informative. This behavior supports the conclusion that the model is not relying primarily on obvious confounders outside the vascular territory.

**Figure 4.**
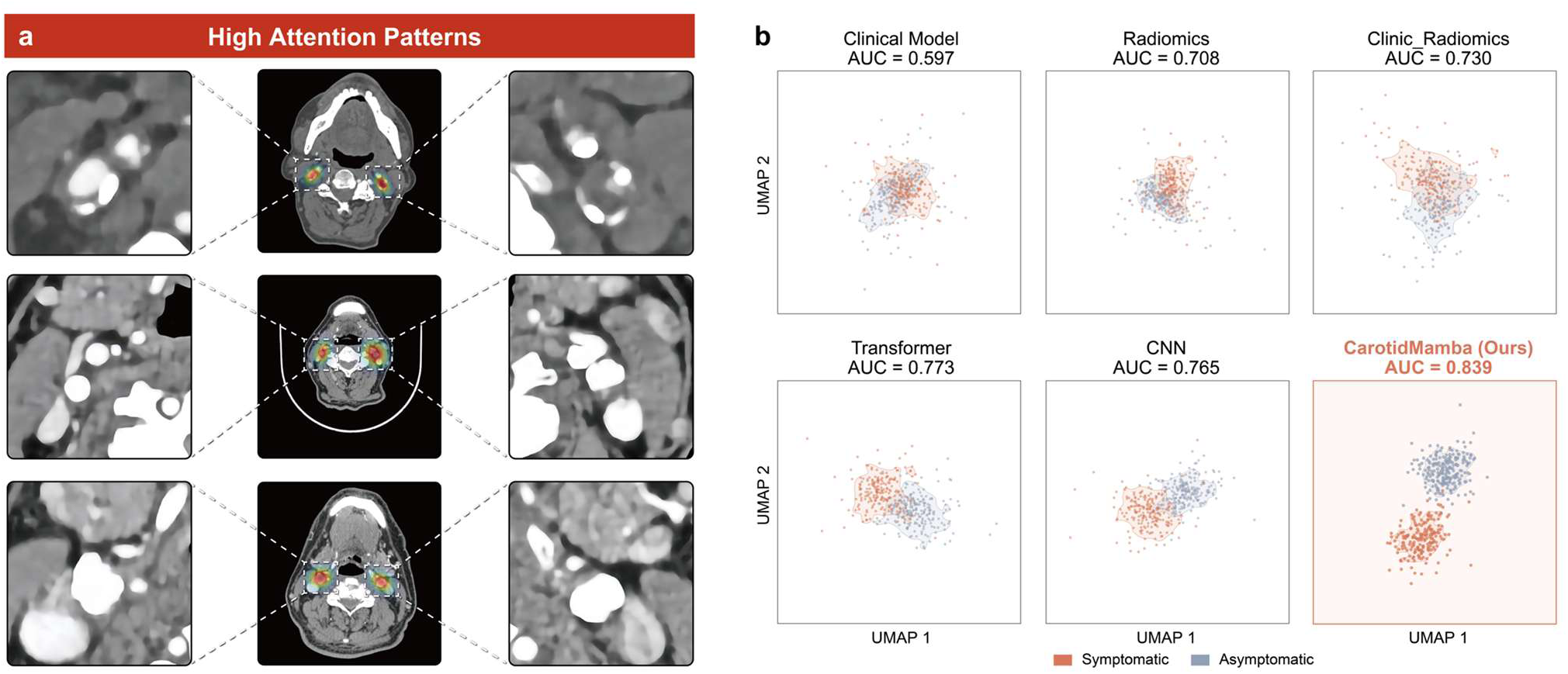
Interpretability and latent-space organization. a, Representative high-attention patterns showing model focus on plaque-bearing carotid regions across axial and reformatted CTA views. b, UMAP projections of the learned feature spaces or output embeddings for the clinical, radiomics, clinic-radiomics, transformer, CNN, and CarotidMamba models. CarotidMamba shows the clearest separation between symptomatic and asymptomatic cases.

We next examined the organization of the learned representation space. UMAP projections of the development-cohort embeddings showed limited separation for the clinical model (AUC, 0.597), modest separation for radiomics (0.708) and clinic-radiomics (0.730), and incremental gains for CNN (0.765) and transformer (0.773). By contrast, CarotidMamba produced the clearest symptomatic–asymptomatic separation, consistent with its development-cohort AUC of 0.839 (Fig. 4b). Together, the saliency and embedding analyses suggest that the model learns a clinically coherent representation of symptomatic plaque phenotype.

## Discussion

In this multi-institutional retrospective CTA study, we developed and validated CarotidMamba, a clinically oriented deep learning framework for phenotyping symptomatic carotid atherosclerotic disease. CarotidMamba combined dual medical foundation models with Mamba-based sequence modeling and achieved stronger discrimination, calibration, and decision-curve utility than clinical, radiomics, clinic-radiomics, CNN, and transformer baselines. Its performance remained robust across three independent external cohorts, and its attention maps and latent-space organization were consistent with biologically plausible plaque localization and class separation.

The main clinical contribution of this work is to support vulnerability-aware carotid CTA assessment beyond stenosis alone. Contemporary carotid management still relies heavily on luminal narrowing thresholds, yet accumulating evidence shows that intraplaque hemorrhage, ulceration, lipid-rich components, and other plaque features carry substantial prognostic information [4–9]. CTA is already embedded in routine stroke workflows, making it a pragmatic substrate for scalable plaque phenotyping [5,6,10,11]. Prior CTA studies have shown that radiomics or machine-learning models can identify symptomatic or high-risk plaques [12,15–20], but most have depended on manually delineated plaque regions, handcrafted features, or single-center development. In contrast, our study evaluated a whole-scan framework in a multi-institutional cohort with three independent external validation datasets. This matters clinically because tools intended to support carotid decision-making must be robust to differences in patient mix, workflow, and imaging environment, not only to a single development center.

From a technical perspective, the observed gains appear to arise from three complementary design choices. First, the dual foundation-model strategy couples disease-aware visual semantics from MedCLIP with anatomy-aware priors from SAM-Med2D, enabling richer slice-level representations than either encoder alone [21–23]. Second, the Mamba encoder is well matched to the ordered nature of CTA, where clinically important evidence may be distributed across multiple nonadjacent slices. Compared with CNN aggregation, this improves long-range context modeling; compared with transformers, it offers a more computationally efficient alternative for long image sequences [24–26,29]. Third, the sequence-rearrangement branch and attention pooling appear to refine how information is integrated across the scan, as supported by the ablation results. Together, these findings help explain why CarotidMamba outperformed both conventional CNN and transformer baselines in internal and external validation.

Interpretability is an important strength of the present framework. Attention maps consistently localized to plaque-bearing carotid territories rather than to unrelated structures, which is particularly relevant in a task that could influence invasive treatment decisions. The UMAP analysis provided a complementary view, showing that the learned representation space more clearly separated symptomatic from asymptomatic cases than the comparator models. Beyond visual explainability, calibration and decision-curve analysis are also clinically important forms of interpretability. Lower Brier scores, lower expected calibration error, and higher net benefit suggest that CarotidMamba is not only ranking patients better, but also producing better behaved probabilities that may be more useful for downstream clinical triage and shared decision-making.

This study has several limitations. First, it was retrospective, and the label captured symptomatic status rather than prospective stroke occurrence; the model therefore identifies a clinically relevant vulnerability-associated phenotype but does not directly predict future events. Second, although external validation was multi-institutional, some cohorts—particularly GH-NTC—were relatively small, and broader temporal and geographic validation remains necessary. Third, the model was trained with patient-level labels rather than lesion-level or side-resolved labels, which may underuse lateralized information in patients with bilateral disease. Future work should therefore emphasize prospective validation, side-resolved phenotyping, richer multimodal integration with laboratory or ultrasound/MR data, and formal reader-assistance studies that quantify clinical impact.

In conclusion, CarotidMamba provides an interpretable and externally validated CTA phenotyping framework for symptomatic carotid atherosclerotic disease. By combining complementary foundation-model representations with efficient Mamba-based sequence modeling, it offers a promising route toward clinically scalable carotid vulnerability assessment beyond stenosis alone.

## Methods

### Study design and cohorts

This retrospective multicenter study included head-and-neck CTA examinations during Jan 2022 – Jan 2026 from four hospitals in China: the First Affiliated Hospital of Dalian Medical University (FAH-DMU), Yingkou Central Hospital (YCH), Dandong Central Hospital (DCH), and the General Hospital of Northern Theater Command (GH-NTC). A total of 705 examinations were screened, and 689 eligible patients were retained after exclusion for incomplete clinical data or poor image quality. The development cohort comprised 423 patients from FAH-DMU, whereas the external validation cohorts comprised 115 patients from YCH, 90 from DCH, and 61 from GH-NTC. The study protocol was reviewed by the ethics committee of the coordinating center (FAH-DMU; approval no. PJ-KS-KY-2026-138) and by participating-site committees (DCH; DDSZXYY-2026-11; YCH; ykzxyy1120260003; GH-NTC:2026-54). Data handling followed local institutional requirements for retrospective, noninterventional research using existing clinical imaging and structured clinical records. The requirement for written informed consent was waived for retrospective research using routinely acquired data.

### Patient eligibility and label definition

Adults 18 years of age or older with bilateral carotid plaques identified on CTA were eligible for inclusion. Carotid plaque was defined according to the Mannheim consensus criteria. Patients were excluded for possible cardiogenic embolism, coagulation disorders, severe comorbidity, estimated renal clearance below 60 mL/min/1.73 m2, allergy to iodinated contrast media, or other neurological disorders such as demyelinating disease or brain tumor.

Symptomatic status was defined as CTA performed within 6 months of a carotid-territory ischemic event, including ischemic stroke or transient ischemic attack, accompanied by manifestations such as hemiplegia, sensory loss, dysarthria, dysphagia, or monocular blindness. Patients scanned 6 months or more after symptom onset were classified as asymptomatic.

### CTA acquisition

Detailed CTA acquisition parameters were available from the coordinating-center protocol. At FAH-DMU, examinations were performed on a Philips Brilliance 64-slice CTA system. Patients were positioned supine with the head tilted backward to reduce dental artifacts. Iodinated contrast (320 mgI/mL) was administered through an antecubital vein using a dual-channel injector at 4.5 mL/s for a total dose of 95 mL. Bolus tracking was performed in the ascending aorta with a 250-HU threshold, with monitoring beginning 10 s after contrast injection and scanning automatically triggered after a 2.2-s delay once the threshold was reached.

Scanning extended from the aortic arch to 2 cm below the cranial vertex. Lead-site acquisition settings were 120 kV tube voltage, 200 mAs tube current, 16 × 0.75 collimation, pitch 1.188, gantry rotation time 0.4 s, matrix size 512 × 512, slice thickness 0.625 mm, and slice interval 0.6 mm. External centers followed site-specific routine CTA protocols that yielded diagnostically comparable axial reconstructions for model analysis.

### Image preprocessing

DICOM data were converted to Hounsfield-unit representations, processed using a vascular window, normalized, and converted into RGB-compatible tensors for feature extraction by the foundation encoders. Each examination was represented as an ordered sequence of 2D CTA slices spanning the carotid territory. No manual lesion-level or slice-level labels were used during model training; instead, patient-level labels were assigned to the full scan, following a weakly supervised instance-aggregation paradigm [27,28].

### CarotidMamba architecture

CarotidMamba uses two pretrained medical foundation encoders to derive complementary slice-level representations (Fig. 1c). For slice *i*, MedCLIP generates a disease-aware feature vector m_i_ and SAM-Med2D generates an anatomy-aware feature vector s_i_. These features are fused by concatenation, z_i_=[m_i_;s_i_], and arranged as an ordered scan-level sequence Z=z_1_,…,z_n_.

The sequence encoder is based on the Mamba family of selective state-space models. The fused slice sequence is linearly projected and processed by stacked Radiology Mamba blocks. A sequence-rearrangement branch provides an alternative ordered view of the same scan to capture complementary long-range dependencies before the sequence is restored. Final patient-level aggregation is performed with attention pooling [27], H=Σ_i_α_i_h_i_, where α_i_=softmax(w^T^tanh(Vh_i_)), and *H* is passed to a classification head for symptomatic versus asymptomatic prediction.

### Comparator models

Five comparator families were evaluated on the same cohort partitions used for CarotidMamba: a clinical model, a radiomics model, a clinic-radiomics model, a CNN baseline, and a transformer baseline. The clinical model was based on logistic regression using structured variables from the FAH-DMU development cohort. The radiomics and clinic-radiomics comparators followed the standard workflow, including region-based handcrafted feature extraction, feature selection, and machine-learning classification. The CNN and transformer comparators used the same scan-level input organization and training partitions as CarotidMamba but replaced the sequence modeling strategy with conventional convolutional or self-attention-based backbones, respectively.

### Training and validation

The FAH-DMU cohort was divided by stratified five-fold cross-validation. In each round, four folds were used for training and one fold for internal validation, yielding five internal performance estimates. The fold-specific models were then evaluated on YCH, DCH, and GH-NTC to obtain external-validation performance under a consistent multi-model protocol.

Training used a Lookahead optimizer with an initial learning rate of 2 × 10−3, weight decay of 1 × 10−4, batch size of one scan per iteration, and cross-entropy loss. Unless otherwise specified, the default CarotidMamba encoder used two Radiology Mamba blocks. Experiments were run on an NVIDIA RTX 4090 GPU.

### Evaluation, calibration, and interpretability analyses

Primary discrimination metrics were AUC, accuracy, precision, F1 score, and recall. Calibration [31] was summarized with the Brier score, expected calibration error, and calibration slope. Clinical utility was assessed by decision-curve [30] net benefit. Confusion matrices were used to visualize cohort-specific error patterns.

Interpretability was examined in two ways. First, representative attention or saliency maps were generated to localize the image regions contributing most strongly to prediction. Second, UMAP [32] was used to project learned representations into two dimensions for visual assessment of symptomatic–asymptomatic separation across methods. Ablation analyses removed or modified individual architectural components, including the Mamba layers, attention pooling, positional encoding, dual-encoder features, sequence rearrangement, encoder depth, aggregation head, and fusion strategy.

## Statistical analysis

Between-group comparisons were performed using the Mann–Whitney U test for continuous variables and the χ² test or Fisher’s exact test for categorical variables, as appropriate. Univariable and multivariable logistic regression analyses were conducted to evaluate the associations between structured clinical factors and symptomatic status in the development cohort and to develop the clinical model. Model performance metrics are reported with 95% confidence intervals. In addition, bootstrap resampling with 1,000 iterations was performed to estimate the variability of model performance and to assess the statistical significance of differences between methods.

## Acknowledgements

This work was supported by the Scientific Research Fund of the Liaoning Provincial Education Department (Grant No. LJKMZ20221280, awarded to GN-J). The authors also gratefully acknowledge financial support from the Hainan Hongji Medical Development Foundation (Grant No. 2025HZ136).

## Data availability

The de-identified CTA images and structured clinical data are not publicly available because of institutional ethics and privacy restrictions. They may be available from the corresponding author upon reasonable request, subject to approval by the participating institutions and an appropriate data-use agreement.

## Code availability

The core CarotidMamba training and inference code will be made available through a version-controlled public repository upon publication when permitted by institutional policy. Because released code cannot include patient-identifiable imaging data, pretrained weights and derived feature files may be shared by the corresponding author upon reasonable request and subject to institutional and ethics approval.

## Author contributions

Y.-S.L., X.-W.D., P.-Y.Z., and G.-N.J. conceived and designed the study. Y.-S.L., X.-W.D., P.-Y.Z., L.-J.M., Y.-N.Y., G.-W.S., J.-G.S., X.Y., F.S., B.-W.Z., and Z.-W.L. curated clinical and imaging data across sites. C.Q., Z.-W.C., X.-H.Q., and X.Y. performed model development, statistical analyses, and figure preparation. Y.-S.L., X.-W.D., and P.-Y.Z. drafted the manuscript. G.-N.J. supervised the study.

## Abbreviations

CTA: computed tomography angiography
AI: artificial intelligence
CNNs: convolutional neural networks
FAH-DMU: the First Affiliated Hospital of Dalian Medical University
YCH: Yingkou Central Hospital
DCH: Dandong Central Hospital
GH-NTC: the General Hospital of Northern Theater Command
OR: odds ratio

## References

1. GBD 2021 Stroke Risk Factor Collaborators. Global, regional, and national burden of stroke and its risk factors, 1990–2021: a systematic analysis for the Global Burden of Disease Study 2021. Lancet Neurol. 2024;23:973–1003. doi:10.1016/S1474-4422(24)00369-7.

2. Saini V, Guada L, Yavagal DR. Global epidemiology of stroke and access to acute ischemic stroke interventions. Neurology. 2021;97(Suppl 2):S6–S16. doi:10.1212/WNL.0000000000012781.

3. Bonati LH, Kakkos S, Berkefeld J, et al. European Stroke Organisation guideline on endarterectomy and stenting for carotid artery stenosis. Eur Stroke J. 2021;6:I–XLVII. doi:10.1177/23969873211012121.

4. Schindler A, Wengier A, Golsari A, et al. Prediction of stroke risk by detection of hemorrhage in carotid plaques: meta-analysis of individual patient data. JACC Cardiovasc Imaging. 2020;13:395–406. doi:10.1016/j.jcmg.2019.03.028.

5. Saba L, Yuan C, Hatsukami TS, et al. Imaging biomarkers of vulnerable carotid plaques for stroke risk prediction and their potential clinical implications. Lancet Neurol. 2019;18:559–572. doi:10.1016/S1474-4422(19)30035-3.

6. Baradaran H, Patel P, Gialdini G, et al. Optimal carotid plaque features on computed tomography angiography associated with ischemic stroke. J Am Heart Assoc. 2021;10:e019462. doi:10.1161/JAHA.120.019462.

7. van Dam-Nolen DHK, van der Kolk AG, de Borst GJ, et al. Carotid plaque characteristics predict recurrent ischemic stroke and transient ischemic attack: the PARISK study. JACC Cardiovasc Imaging. 2022;15:1715–1726. doi:10.1016/j.jcmg.2022.04.003.

8. Waksman R, Torguson R. The vulnerable plaque detected: time to consider treatment. Lancet. 2021;397:943–945. doi:10.1016/S0140-6736(21)00504-3.

9. Derdeyn CP, Chaturvedi S, Wasserman BA. Shrinking role for carotid revascularization in stroke prevention. Stroke. 2025;56:2830–2833. doi:10.1161/STROKEAHA.125.051981.

10. Kolossváry M, De Cecco CN, Feuchtner G, Maurovich-Horvat P. Advanced atherosclerosis imaging by CT: radiomics, machine learning and deep learning. J Cardiovasc Comput Tomogr. 2019;13:274–280. doi:10.1016/j.jcct.2019.04.007.

11. Zaccagna F, Rundo L, Puglisi S, et al. CT texture-based radiomics analysis of carotid arteries identifies vulnerable patients: a preliminary outcome study. Neuroradiology. 2021;63:1043–1052. doi:10.1007/s00234-020-02628-0.

12. Pisu F, Saba L, Balestrino A, et al. Machine learning detects symptomatic plaques in patients with carotid atherosclerosis on CT angiography. Circ Cardiovasc Imaging. 2024;17:e016274. doi:10.1161/CIRCIMAGING.123.016274.

13. Zhu Y, Chen L, Lu W, Gong Y, Wang X. The application of the nnU-Net-based automatic segmentation model in assisting carotid artery stenosis and carotid atherosclerotic plaque evaluation. Front Physiol. 2022;13:1057800. doi:10.3389/fphys.2022.1057800.

14. Zhai D, Liu R, Liu Y, et al. Deep learning-based fully automatic screening of carotid artery plaques in computed tomography angiography: a multicenter study. Clin Radiol. 2024;79:e994–e1002. doi:10.1016/j.crad.2024.04.015.

15. Liu M, Chang N, Zhang S, et al. Identification of vulnerable carotid plaque with CT-based radiomics nomogram. Clin Radiol. 2023;78:e856–e863. doi:10.1016/j.crad.2023.07.018.

16. Chen C, Tang W, Chen Y, et al. Computed tomography angiography-based radiomics model to identify high-risk carotid plaques. Quant Imaging Med Surg. 2023;13:6089–6104. doi:10.21037/qims-23-158.

17. Shi J, Sun Y, Hou J, et al. Radiomics signatures of carotid plaque on computed tomography angiography: an approach to identify symptomatic plaques. Clin Neuroradiol. 2023;33:931–941. doi:10.1007/s00062-023-01289-9.

18. Nie JY, Chen WX, Zhu Z, et al. Initial experience with radiomics of carotid perivascular adipose tissue in identifying symptomatic plaque. Front Neurol. 2024;15:1340202. doi:10.3389/fneur.2024.1340202.

19. Yang Y, Wei J, Wei X, et al. Interpretable machine learning for detecting symptomatic patients with carotid atherosclerosis on computed tomography angiography: a retrospective diagnostic study. BMC Med Imaging. 2025;25:521. doi:10.1186/s12880-025-02113-1.

20. Wang H, Xu J, Ye C, et al. An explainable CT-based machine learning model integrating carotid plaque and perivascular adipose tissue for predicting symptomatic plaques. Front Neurol. 2025;16:1679861. doi:10.3389/fneur.2025.1679861.

21. Wang Z, Wu Z, Agarwal D, Sun J. MedCLIP: contrastive learning from unpaired medical images and text. In: Proceedings of the 2022 Conference on Empirical Methods in Natural Language Processing. Association for Computational Linguistics; 2022:3876–3887.

22. Kirillov A, Mintun E, Ravi N, et al. Segment Anything. In: Proceedings of the IEEE/CVF International Conference on Computer Vision. 2023:4015–4026.

23. Ma J, He Y, Li F, et al. Segment anything in medical images. Nat Commun. 2024;15:654. doi:10.1038/s41467-024-44824-z.

24. Gu A, Goel K, Ré C. Efficiently modeling long sequences with structured state spaces. In: International Conference on Learning Representations. 2022.

25. Gu A, Dao T. Mamba: linear-time sequence modeling with selective state spaces. In: First Conference on Language Modeling. 2024.

26. Yang S, Wang Y, Chen H. MambaMIL: enhancing long sequence modeling with sequence reordering in computational pathology. In: Medical Image Computing and Computer-Assisted Intervention—MICCAI 2024. Springer; 2024:296–306.

27. Ilse M, Tomczak JM, Welling M. Attention-based deep multiple instance learning. In: Proceedings of the 35th International Conference on Machine Learning. PMLR; 2018;80:2127–2136.

28. Lu MY, Williamson DFK, Chen TY, et al. Data-efficient and weakly supervised computational pathology on whole-slide images. Nat Biomed Eng. 2021;5:555–570. doi:10.1038/s41551-020-00682-w.

29. Shao Z, Bian H, Chen Y, et al. TransMIL: transformer-based correlated multiple instance learning for whole slide image classification. Adv Neural Inf Process Syst. 2021;34:2136–2147.

30. Vickers AJ, Elkin EB. Decision curve analysis: a novel method for evaluating prediction models. Med Decis Making. 2006;26:565–574. doi:10.1177/0272989X06295361.

31. Steyerberg EW, Vickers AJ, Cook NR, et al. Assessing the performance of prediction models: a framework for traditional and novel measures. Epidemiology. 2010;21:128–138. doi:10.1097/EDE.0b013e3181c30fb2.

32. McInnes L, Healy J, Melville J. UMAP: uniform manifold approximation and projection for dimension reduction. arXiv. 2018;1802.03426.

